# Prebiotic Supplements Correct Oral Probiotic Deficiency for Lasting Allergy Relief

**DOI:** 10.1101/2023.07.20.23291433

**Authors:** Cliff Shunsheng Han

## Abstract

Allergic rhinitis has increased in the last fifty years from affecting less than a percent to more than twenty-six percent of the population. Today, more than one hundred million people in the US suffer from these seasonal or yearlong allergies. The hygiene hypothesis was proposed 30 years ago as a potential explanation for this phenomenon, and we built on that with the specific oral hygiene hypothesis. Our longitudinal pilot study suggested that oral probiotic deficiency is the cause of allergic rhinitis. This clinical trial served to verify our theory and evaluate the effectiveness of AllerPops for allergy relief. Through it, we show that AllerPops prebiotic supplements are effective in providing sustained allergy relief (p = 0.002) and can modulate oral beneficial bacteria that produce short-chain fatty acids (SCFA), such as *Fusobacteria, Butyrivibrio*, and *Peptostreptococcus*. The clinical improvements correlated with changes in the relative abundance of probiotics significantly: *Fusobacteria* (R = 0.32, p = 0.009), *Butyrivibrio* (R = 0.25, p = 0.044), and *Peptostreptococcus* (R = 0.34, p = 0.005). These results point to the root cause of allergic rhinitis: the lack of oral probiotics that produce SCFA to pacify the immune systems. Future study of AllerPops’ theory will help society redefine the best oral hygiene practice to protect oral probiotics so that we may prevent allergic and autoimmune diseases and dental/gum infections. The trial was retrospectively registered at clinicaltrials.com, with registration number NCT05956691, on 21/07/2023.

---

Allergic rhinitis (AR) was described in 1929 as a process that includes three cardinal symptoms: sneezing, nasal obstruction, and mucus discharge[1]. AR is clinically defined as a symptomatic disorder of the nose induced by an immunoglobulin(IgE)-mediated inflammation after allergen exposure of the membranes lining the nose. AR is a widely prevalent condition in Canada (i.e., 20 to 25%)[2] and the United States[3]. Significant physical sequelae and recurrent or persistent morbidities often accompany it.

More than 30 years ago, Dr. Strachan proposed the hygiene hypothesis to explain the increase in allergy diseases[4]. Subsequently, many later studies suggest that personal and social hygiene practices are associated with the allergy disease epidemic[5]. Increasing literature on animals and humans has implicated changes in the gut microbiome associated with the development of allergic disease[6-9], which were shown to be associated with reduced early microbial exposure[10]. Changes in the nasal[11-15] and oral microbiota[16-19] were also found.

Some investigations have shown that the oral administration of probiotics and prebiotics may benefit allergic rhinitis patients[20-22]. The local nasal administration of *Lactococcus lactis* NZ9000 can affect the local and systemic immune responses against *Streptococcus pneumonia*[23].

Probiotics are a promising therapeutic approach for allergic airway diseases. However, attempts at using probiotics to cure or prevent allergy diseases have had limited success[24]. In those trials, researchers used probiotics that live in the gut. Instead of focusing on the remote interaction between the gut and the airway, we examined the local interaction between microbiota living in the airway, especially in the mouth, and the immune system residing around the respiratory tracts in our pilot study[25]. As a result, that study illustrates that restructuring oral microbiota can lead to lasting remissions of common allergies (allergic rhinitis).

The primary purpose of this clinical study was to assess the effectiveness of AllerPops[26] in relieving nasal symptoms in volunteers with seasonal or year-long allergies, as indicated by results on the total nasal symptoms score (TNSS) questionnaire and peak nasal inspiratory flow (PNIF) measurement device. The secondary purpose was to identify the changes in the oral microbiome during the intervention and to verify the oral probiotic deficiency hypothesis, that the lack of bacteria that produce SCFA causes allergic rhinitis.

## RESULTS

### Demographics & Groups

Seventy-two (72) individuals clinically diagnosed with persistent or intermittent allergic rhinitis were enrolled randomly into the two arms of the double-blinded phase II clinical trial. Two participants in the experimental treatment arm did not complete the study. Both participants withdrew from the study after baseline values were taken (CONSORT Diagram, Supplementary Figure S1).

Participants included 23 males and 49 females, with an average age of 37.8 years +/- SD 14.1 (range 18 – 70), a height of 168.9 cm +/- SD 9.6 (range 147.5 – 197.0), a weight of 68.7 kg +/- SD 12.2 (range 47.3 – 106.9), and a BMI of 24.0 kg/m2 SD 3.1 (range 18.6 – 29.9).

Participants were randomized to either the control group or the investigational group. No statistically significant differences between the control group (n = 36) and the investigational group (n = 36) were seen in any of the baseline measurements aside from body temperature (See Table 1); the slight mean difference of 0.15°C can be explained by more females being enrolled in the control group.

**Table 1.** Demographics and baseline measurements for the control and investigational groups.

| Measurement | Control Group (*n* = 36) | Investigational Group (*n* = 36) | p-value |
| --- | --- | --- | --- |
| Age (*mean, SD, in years*) | 39.4 (13.9) | 35.3 (14.3) | 0.11 |
| Height (*mean, SD, in cm*) | 169.7 (10) | 167.8 (9.4) | 0.40 |
| Weight (*mean, SD, in kg*) | 70 (13.2) | 67.6 (11.7) | 0.38 |
| BMI (*mean, SD, kg/m<sup>2</sup>*) | 24.2 (3.1) | 23.9 (3.2) | 0.72 |
| Female | 69.4% | 66.7% | 0.84 |
| TNSS (*mean, SD*) |  |  |  |
| *Nasal Obstruction* | 1.8 (0.62) | 1.5 (0.65) | 0.10 |
| *Itching/Sneezing* | 1.4 (0.77) | 1.4 (0.73) | 0.98 |
| *Secretion/Runny Nose* | 1.5 (0.91) | 1.4 (0.81) | 0.66 |
| *Overall* | 4.8 (1.92) | 4.4 (1.78) | 0.46 |
| PNIF (*mean, SD, in L/min*) | 72.8 (32.2) | 79.5 (37.2) | 0.41 |
| IgE (*mean, SD, IU/mL*) | 121.7 (154.3) | 140.7 (191.6) | 0.99 |
| Eosinophils (*mean, SD, x10<sup>9</sup>/L*) | 0.18 (0.17) | 0.21 (0.15) | 0.43 |
| Heart Rate (*mean, SD, beats/min*) | 72.4 (10.8) | 74.4 (10) | 0.30 |
| Body Temperature (*mean, SD, °C*) | 36.3 (0.7) | 36.2 (0.6) | **0.046** |
| Blood Pressure (*mean, SD, mm Hg*) |  |  |  |
| *Systolic* | 118.1 (12) | 116.9 (9.8) | 0.73 |
| *Diastolic* | 72.7 (7.4) | 73.2 (8.2) | 0.58 |

### Allerpops Relieves Allergies according to TNSS and PNIF Score

Baseline TNSS scores were obtained for all participants, with overall scores averaging 4.6 +/- SD 1.8, based on the following breakdown:

- nasal obstruction score of 1.7 +/- SD 0.7,
- itchiness score of 1.4 +/- SD 0.7, and
- runny nose score of 1.5 +/- SD 0.9.

PNIF measurements averaged 76.1 L/min +/- SD 34.7. A study by Dor-Wojnarowska in 2011 established reference PNIF values. Participants in this study had a mean PNIF value 57.7 L/min lower than they were expected to have given their sex and height (i.e., 133.8 L/min), a difference anticipated with all participants having been clinically diagnosed with allergic rhinitis.

To assess treatment impact, the change in TNSS scores was compared between the control and investigational groups (See Table 2). Each group improved on both the overall score and the three sub-scores during all three assessments (i.e., day 7, day 14, and day 21), with this change only being statistically significant between the two groups on day 14 for the overall score (p = 0.016) and the nasal sub-score (p = 0.032).

**Table 2.** Change in TNSS, PNIF values, eosinophils, and IgE from baseline in the control and investigational groups on days 7, 14, and 21.

| Measurement | Control Group (n = 33 to 35) |  |  | Investigational Group (n = 30 to 34) |  |  | p-value |  |  |
| --- | --- | --- | --- | --- | --- | --- | --- | --- | --- |
|  | Day 7 | Day 14 | Day 21 | Day 7 | Day 14 | Day 21 | Day 7 | Day 14 | Day 21 |
| TNSS (change, SD) |  |  |  |  |  |  |  |  |  |
| Nasal Obstruction | -0.6 (0.7) | -0.6 (0.8) | -0.7 (0.9) | -0.3 (0.8) | -0.2 (0.8) | -0.3 (1) | 0.17 | 0.032 | 0.08 |
| Itching/Sneezing | -0.4 (0.8) | -0.7 (0.9) | -0.6 (0.9) | -0.2 (0.7) | -0.4 (0.8) | -0.5 (1.1) | 0.67 | 0.11 | 0.71 |
| Secretion/Runny Nose | -0.5 (0.9) | -0.7 (0.8) | -0.7 (0.8) | -0.1 (0.8) | -0.4 (0.9) | -0.3 (0.8) | 0.22 | 0.10 | 0.053 |
| OVERALL | -1.4 (2) | -2.1 (1.9) | -2 (2) | -0.6 (1.8) | -1.1 (2) | -1.1 (2.6) | 0.19 | 0.016 | 0.079 |
| PNIF (change, SD, in L/min) | 18.1 (38) | 29 (41.5) | 17.8 (24.2) | 6.7 (42.6) | 4.4 (42.1) | 8.7 (45.2) | 0.38 | 0.045 | 0.14 |
| Eosinophils (mean, SD, x10<sup>9</sup>/L) | 0.003 (0.05) | -- | -0.006 (0.07) | 0.004 (0.04) | -- | -0.02 (0.07) | 0.99 | -- | 0.73 |
| IgE (change, SD, IU/mL) | 2.2 (32.8) | -- | -17.3 (134.2) | 1.4 (17.5) | -- | 4.6 (31.6) | 0.62 | -- | 0.80 |

PNIF scores also improved in both groups, although the difference between the two groups was only statistically significant on the day 14 mark (p = 0.045) (See Table 2).

The overall TNSS score for the investigational group improved significantly, decreasing by 1.1 through day 14 (p-value = 0.002) (See Figure 1 and Table 3). The overall change was also significant for the runny nose (p = 0.022) and itching/sneezing (p = 0.002) TNSS sub-scores. Analysis with the Wilcoxon signed-rank test found all three changes above to be significantly different from baseline at the 14-day mark (p-values ranging from 0.004 to 0.013), with one change (i.e., overall) being significant at day 7 (p = 0.033), and two changes (i.e., overall and itching/sneezing) being significant at 21 days (p = 0.017 and 0.025). TNSS also improved significantly in the control group overall and for all three sub-scores (p < 0.00001 for all four changes). When comparing TNSS changes week-over-week against baseline, no significant changes were observed between days 14 to 21 for either the control group (p-values ranging from 0.14 to 1.00) or the investigational group (p-values ranging from 0.27 to 0.67).

**Table 3.**
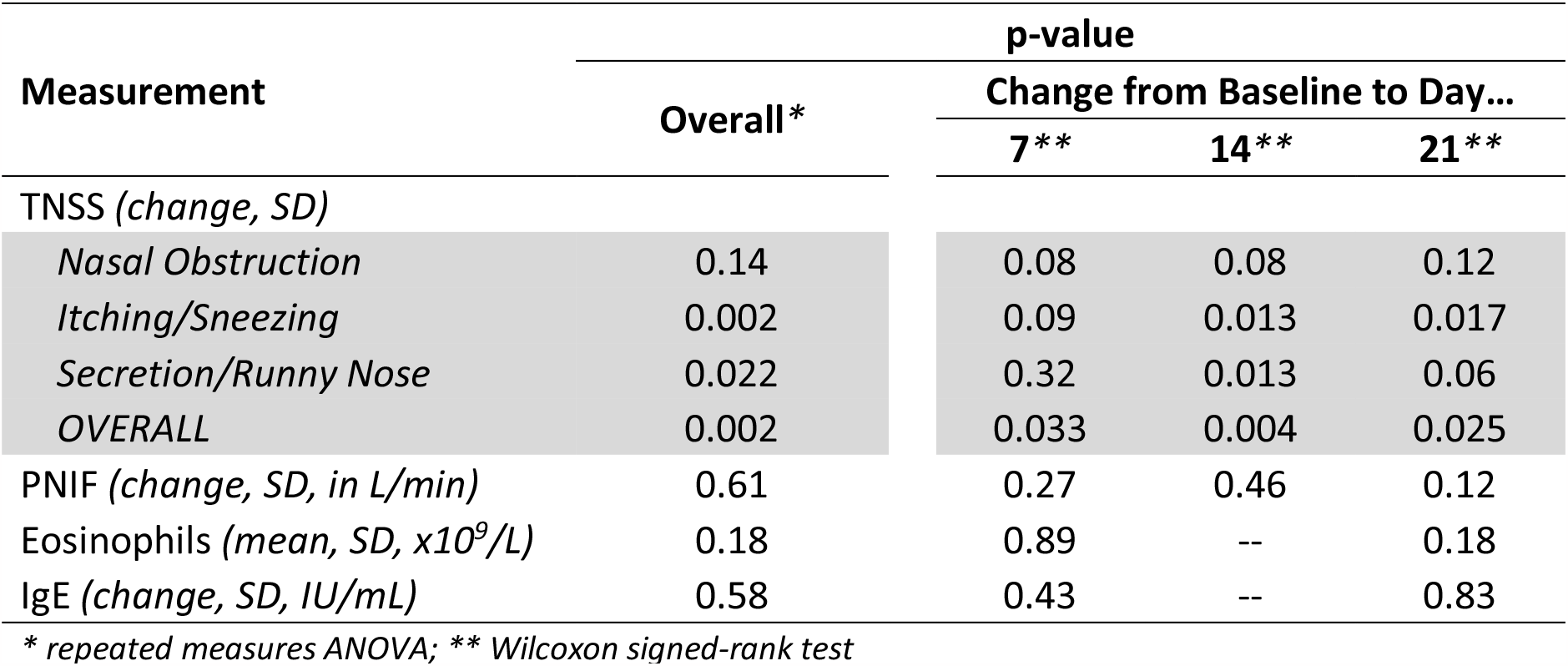
Change in TNSS, PNIF values, eosinophils, and IgE from baseline to day 7/14/21 in the investigational group.

| Measurement | Overall* | p-value |  |  |
| --- | --- | --- | --- | --- |
|  |  | Change from Baseline to Day... |  |  |
|  |  | 7** | 14** | 21** |
| TNSS (change, SD) |  |  |  |  |
| Nasal Obstruction | 0.14 | 0.08 | 0.08 | 0.12 |
| Itching/Sneezing | 0.002 | 0.09 | 0.013 | 0.017 |
| Secretion/Runny Nose | 0.022 | 0.32 | 0.013 | 0.06 |
| OVERALL | 0.002 | 0.033 | 0.004 | 0.025 |
| PNIF (change, SD, in L/min) | 0.61 | 0.27 | 0.46 | 0.12 |
| Eosinophils (mean, SD, x10<sup>9</sup>/L) | 0.18 | 0.89 | -- | 0.18 |
| IgE (change, SD, IU/mL) | 0.58 | 0.43 | -- | 0.83 |

\* repeated measures ANOVA; \*\* Wilcoxon signed-rank test

**Figure 1.**
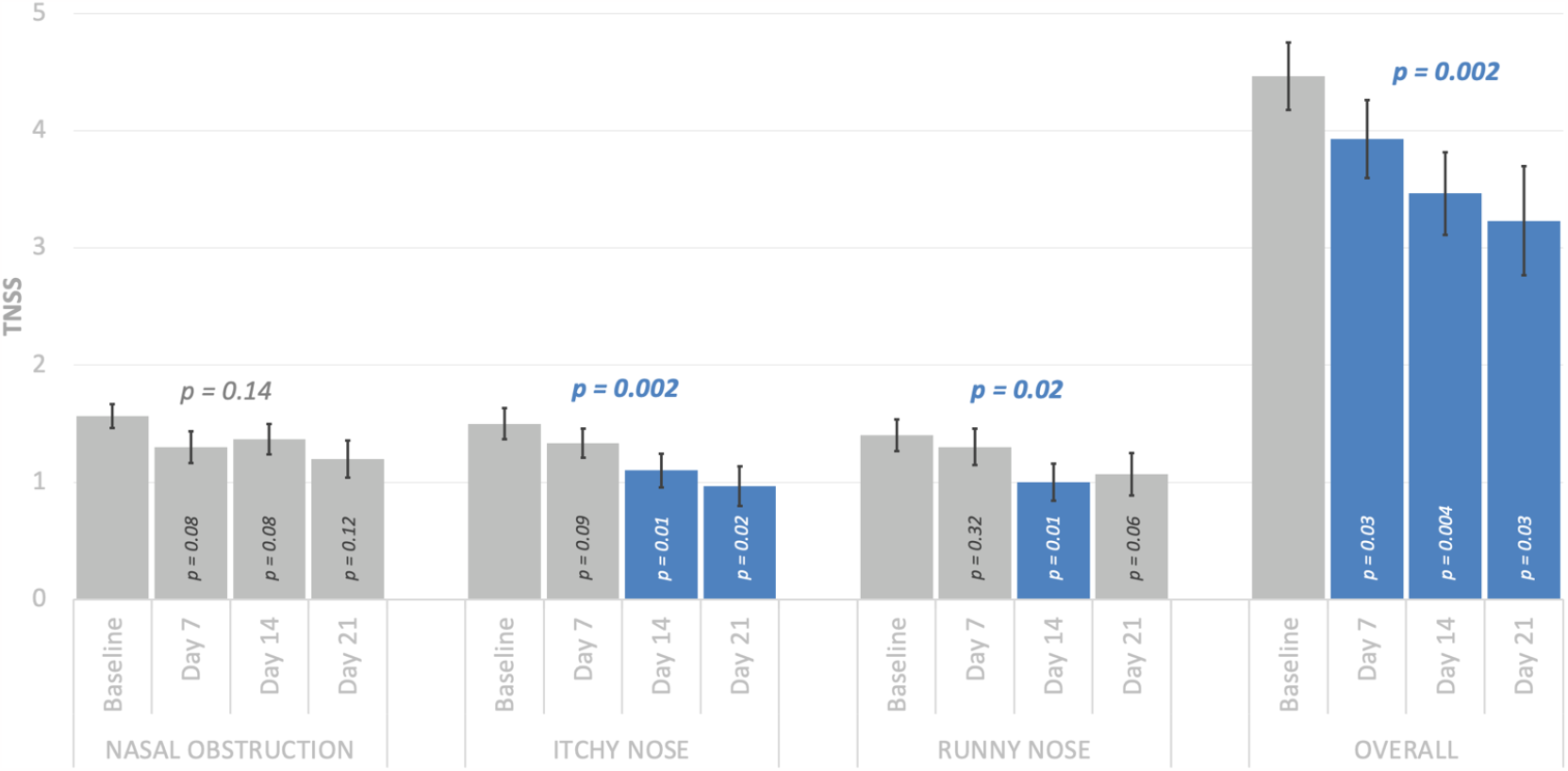
Comparison of TNSS from baseline to 7, 14, and 21 after the start of treatment. Bar represents the mean; whiskers represent positive and negative standard error. Blue text and bars represent statistical significance at the 95% level. The overall p-value was calculated using RM-ANOVA, while the change from baseline for each week was calculated using the Wilcoxon signed-rank test.

Although PNIF values improved for both the investigational and control groups, the change from baseline was significant in the control group (p = 0.00002) but not in the investigational group (p = 0.61). Similar to TNSS changes, PNIF values did not experience a significant change between days 14 to 21 for either the control group (p = 0.40) or the investigational group (p = 0.86).

IgE and eosinophils levels both changed throughout the treatment period but never at a level deemed statistically significant.

### 16S Profiling of Saliva Microbiome Indicates AllerPops Promotes Oral Probiotics

The amplicon was sequenced on Illumina paired-end platform to generate 250 bp paired-end raw reads (Raw PE) and then merged and pre-treated to obtain Clean Tags. The chimeric sequences in Clean Tags were detected and removed to obtain the Effective Tags, which can be used for subsequent analysis. Each sample had a minimum of 100,000 sequences.

Operational Taxonomic Units (OTUs) were obtained by clustering with 97% identity on the Effective Tags of all samples to study the microbial community composition in each sample. Then they identified the level of the kingdom, phyla, class, order, family, genus, and species. Each saliva sample has 215 - 658 observed species, averaging 341 (Supplemental Table S1).

Clustering analysis was used to construct a cluster tree to study the similarity among different samples. The Unweighted Pair-group Method with Arithmetic Mean (UPGMA) is a hierarchical clustering method widely used in ecology to classify samples[27]. Nine participants (eight from the experimental group and one from the control group, Supplemental Figure S2) have their baseline and final samples clustered next to each other, which indicates that the microbiome in the paired samples had minimal changes during the intervention. However, their TNSS score improvements are similar to other subjects (1.77 verse 1.73). This result indicates that changes not detected by this method may contribute the clinical improvement.

The t-test to compare the relative abundance between treatment groups detected in the control group genus *Rothia* decreased after the intervention compared to baseline (from 4.0% to 2.6%, P = 0.024), and genus *unclassified_Clostridia_UCG-014* increased (from 0.1% to 0.2%, P = 0.049). The physiological significance of the change is unknown. Other differences between the two trial groups are likely unrelated to the treatment.

In addition to the standard analysis by Novogene, we performed a correlation test between changes in TNSS score and changes in relative abundance at the genus level. The abundance of three genera is positively correlated with the clinical symptom change (figure 2) and determined to be statistically significant. The three genera are *Fusobacteria* (R = 0.32, p = 0.009), *Butyrivibrio* (R = 0.25, p = 0.044), and *Peptostreptococcus* (R = 0.34, p = 0.005). All three genera can produce short-chain fatty acids[28-30], a group of chemicals that can calm the immune system[31].

**Figure 2.**
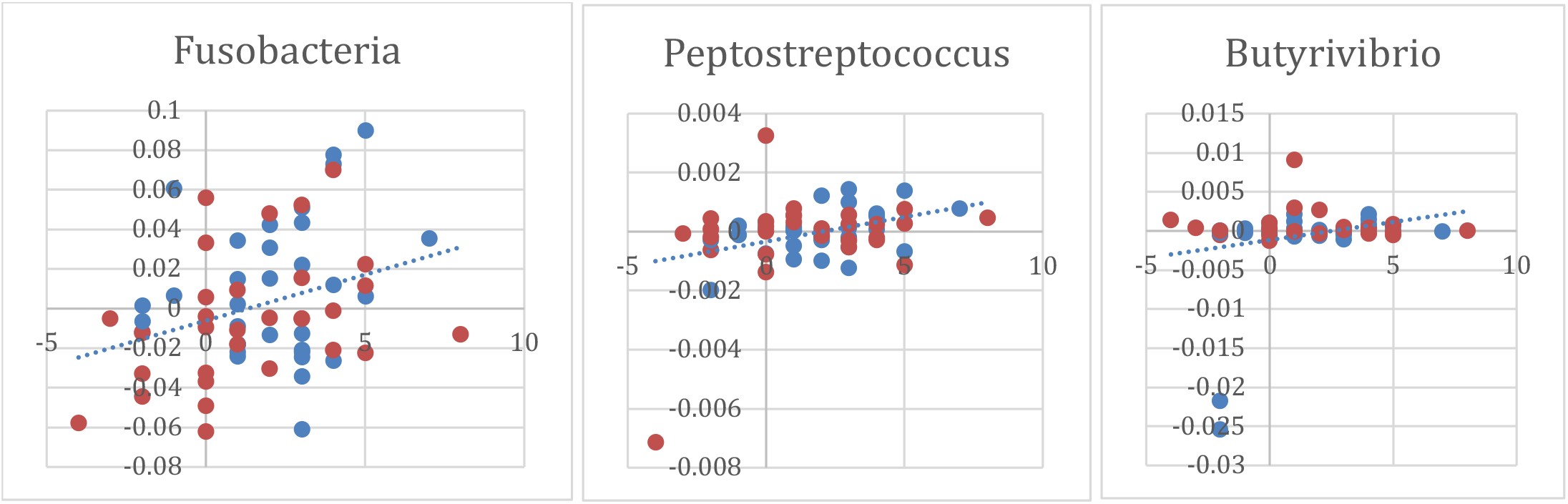
Positive correlation between TNSS score changes (x axle) and relative abundance changes (Y axle) of *Fusobacteria* (R = 0.32, p = 0.009), *Butyrivibrio* (R = 0.25, p = 0.044), and *Peptostreptococcus* (R = 0.34, p = 0.005). Blue, slow dissolving group; Red, fast ingestion group.

On average, the relative abundance of *Fusobacteria* increased by 0.17%, which was not statistically significant. In addition, one outlier for *Peptostreptococcus* and two for *Butyrivibrio* were recorded, with significantly reduced relative abundances. These outliers likely resulted from events unrelated to this trial. The two bacteria’s relative abundance also increased on average for all other participants. Changes in the relative abundance of *Streptococcus* and *Veillonella* previously identified in our pilot study are insignificant in this trial data set.

## DISCUSSION

### AllerPops Prebiotic Supplements Relieve Allergies within a Week

The primary outcome of this clinical trial was to determine the effectiveness of AllerPops in relieving nasal symptoms at the day seven mark and to assess the impact of two different administration methods on the level of relief. Individuals receiving AllerPops as a potential treatment for allergic rhinitis subjectively experienced improvements in nasal symptoms, compared to their baseline value, in both their overall TNSS score and all three TNSS sub-scores.

This improvement occurred in both the control and investigational groups. The improvement in the overall score in the investigational group was statistically significant (p = 0.025), with the change in nasal obstruction showing the most improvement, decreasing by 0.3. Similarly, PNIF values showed an improvement of 28.8% or 6.7 L/min (p = 0.27) over the first seven days of treatment.

### AllerPops Provide Sustained Allergy Relief

A secondary analysis was completed to assess the impact of treatment on symptoms beyond the seven-day mark and into weeks two and three. There was a significant (p = 0.002) positive effect of AllerPops on participant TNSS overall score, as well as for the itching (p = 0.002) and runny nose (p = 0.022) sub-scores. Results, broken down week-by-week, showed that a 14-day intervention period appeared superior, with 75% of sub-scores showing a significant change compared to baseline, and all four scores showing the most significant improvement compared to the 7-day and 21-day intervention periods (See Figure 1).

No significant changes were seen after day 14 in the investigational or control group for any TNSS score. As participants did not consume any product after day 13, this confirms that the initial symptom improvement was strictly due to AllerPops, regardless of how they were consumed. In addition, this finding strongly suggests that both sets of improvements persisted for at least one week after treatment.

In contrast, for PNIF values, while descriptive statistics revealed that participants did improve when slowly consuming AllerPops (i.e., left to melt in the mouth rather than swallowing), this difference was not significant (p = 0.61). For PNIF improvement, the maximum effect happened on day 21, with PNIF values improving by a mean of 8.7 L/min. Also, contrary to TNSS, the slightest change in PNIF values happened on day 14. For the group quickly consuming AllerPops, the change in PNIF values from baseline to day 21 was significant (p = 0.00002), with the change from day 14 to 21 not being significant (p = 0.40). This provides further evidence that improvement was due to AllerPops, regardless of how it was consumed, and that this improvement was sustained for at least one week after the product was last ingested.

However, AllerPops did not reach its full potential in the experiment group due to regulatory restraints. Most significantly, we could not introduce oral hygiene changes in the trial. The extra oral hygiene before and while taking AllerPops may have slowed improvement and negatively impacted the oral probiotics for some participants.

### AllerPops’ Allergy Relief Correlated with Changes in Saliva Microbiome

The microbiome analyses of saliva samples collected from participants before, during, and after the intervention yield both consistent and inconsistent with our pilot study. The inconsistency is that we did not find that two genera, *Streptococcus* and *Veillonella*, became more abundant after using AllerPops. Instead, we found that the variations of three other genera, *Fusobacteria, Butyrivibrio*, and *Peptostreptococcus*, positively correlate with improving clinical symptom scores. These three genera can all produce SCFAs, pacifying the immune system[31]. Therefore, these results are consistent with our previously proposed etiological theory on allergic rhinitis. Oral probiotic deficiency, or the lack of probiotics that produce metabolites, such as SCFA, causes the immune system to become hypersensitive, leading to allergic inflammation[25].

The fact that all three bacterial genera that correlated with the shift of clinical symptoms are SCFA producers further confirms the hypothesis. There are 13 known SCFA producers among the 49 most common genera analyzed. The chance that all three associated general are SCFA producers is 1.55% in this analysis. Insufficient SCFA is likely the major, if not the only, microbe-related mechanism leading to allergies.

The new results indicate that the human immune system and microbiota interaction is very complicated. Multiple bacterial genera participate in the process of calming down the immune system. We previously regarded these bacteria as negative triggers for the human immune system. It is also possible that these bacteria are simply the balancing force against positive triggers, likely MAMPs (microbial-associated molecular pattern) from commensal bacteria. In this and previous studies, we identified five genera as balancing forces of the immune system. Other common oral bacteria, such as genera *Porphyromonas, Prevotella*, and so on [28, 32], that can also produce SCFA likely contribute to the negative control of the immune system.

Each of the thirteen oral SCFA producers may contribute only a tiny portion of the negative control to the immune system in and around the mouth, airway and upper digestion tract. This is likely part of why previous studies have not consistently associated any of the bacteria with allergies [33]. The situation is similar in the gut with many SCFA producing bacteria. Many studies recognize the importance of these groups of gut bacteria in health and preventing diseases[34]. A recent study identified a reduced relative abundance of *Prevotella* and *Veillonella* in the mouth of people with peanut allergies, which is consistent with our observation and supports the hypothesis that food allergies and allergic rhinitis may have a common root cause [35].

These three bacterial genera, *Fusobacteria, Butyrivibrio*, and *Peptostreptococcus*, are part of the oral microbiome in all participants and almost all 205 samples except two, likely due to sampling bias of low abundant genera *Butyrivibrio*. Most of these bacteria are anaerobic, can live in the pocket between the teeth and gums, and have been identified in sites of periodontal disease[36].

On the one hand, these bacteria at sufficient levels can prevent allergies. Conversely, they may worsen things when chronic gum infections occur. Proper oral hygiene is likely the key to keeping these bacteria in check and staying on the good side of allergy prevention with minimal harm to the teeth and gum. Further studies are needed to redefine the boundary of the best practice for oral hygiene for professionals and the general public.

A balance of positive and negative triggers to the immune system is necessary to achieve a healthy state. As Snyder et al.’s germ-free mice experiment demonstrated, a balanced condition can be one with no interaction on either side[37]. We discussed how the balance falters during diseases in our pilot study[25]. Briefly, on one side, acute infection, fever, and diarrhea remove probiotics so that the immune system can fight infectious agents without restraints. This suggests that one should not treat fever and diarrhea prematurely before infections fade. On the other side, chronic infections may arise from probiotic overgrowth.

### Oral Probiotic Deficiency Causes Allergic Rhinitis

We have identified several bacterial genera that likely contribute to initiating allergic rhinitis and have regarded them as probiotics for their capacity to produce SCFA and calm down the immune system. The first critical function of a microbiota associated with a host should be communicating “peace” with the host. Bacteria in this role should (1) not cause immediate harm to the host and (2) be able to send pacifying signals to the host’s immune system. In humans, this communication is likely achieved through SFCA-producing bacteria. These bacteria include *Streptococcus, Veillonella, Fusobacteria, Porphyromonas*, and so on in the mouth and airway, fiber-degrading bacteria in the gut, and *Cutibacterium acnes* on the skin. However, it is vital to remember that these probiotics are not always good and may contribute to pathogenicity in certain disease conditions.

These SCFA-producing probiotics are negative immune system triggers, and removing and reapplying them can be relatively easy and quick. Theoretically, the effects of the negative trigger should not last long after removing it. Otherwise, the immune system’s power cannot be released in time to protect the host. Many different cells in the human body can absorb and metabolize SCFAs, which can rapidly decrease levels of SCFA. Several observations support this proposition. (1) The epidemic of common allergies progresses reasonably quickly in a short time at a population level. (2) Oral biofilm disappears in less than a day under moderate fever. (3) The prebiotic compound promotes oral probiotics to produce fast and lasting allergy relief.

It was hypothesized that immune-mediated disorders such as allergic rhinitis are linked to reduced early microbial exposure, gut dysbiosis, and nasal and oral microbiota. AllerPops contains L-Arginine, which is capable of providing balanced nutritional support and restoration to the oral microbiota, alleviating AR symptoms. The results from this trial and our pilot study link the cause of allergic rhinitis to oral microbiota, the lack of good bacteria, or oral probiotic deficiency, causes allergic rhinitis. Additional testing should be performed to verify this theory on a larger scale.

Overall, this study found AllerPops safe and effective in individuals with allergic rhinitis, whether they swallowed the product or allowed it to melt in their mouth. Oral hygiene requirements in the treatment group and the method of consumption for the control group may have limited the impact and effectiveness of the investigational product.

## SUMMARY

Allergic rhinitis is commonly associated with an itchy, stuffy, or runny nose. Whether consumed quickly or slowly, AllerPops improved allergy symptoms significantly and airflow in the nasal airway. Furthermore, these improvements did not deteriorate one week after investigational product consumption was terminated. AllerPops’s capability to promote oral probiotics enables sustained allergy relief.

## MATERIALS AND METHODS

### STUDY DESIGN

The study was conducted as a phase II, a randomized, double-blind, controlled, single-center, 21-day study designed to investigate the efficacy of AllerPops in reducing nasal symptoms in adult participants with seasonal/year-long nasal allergies. Participants were dosed every other day for a minimum of 3 doses, and after that, until the participant was satisfied with the relief of the nasal allergy symptoms.

#### Ethics statements

All methods were carried out in accordance with relevant guidelines and regulations.All experimental protocols were approved by Health Canada with protocol number 21-SAHSE-01, and Advarra Institutional Review Board (Aurora, ON, Canada). Informed consent was obtained from all subjects before participation.

### PARTICIPANTS

All participants were between 18 and 70 years old and clinically diagnosed with persistent or intermittent AR according to the Allergic Rhinitis and Its Impact on Asthma (ARIA) classification[38]. Participants meeting any exclusion criteria were not eligible for admission to the study. A baseline total nasal symptom score (TNSS) was obtained from all participants. TNSS is a questionnaire developed to assess AR symptoms. In this subjective assessment, AR symptoms were scored by the participant from 0 to 3 (0 = none, 1 = mild, 2 = moderate, 3 = severe)[39]. Peak nasal inspiratory flow (PNIF) scores were also obtained. This device assesses nasal airway patency by measuring nasal volume and flow[40], and provides an objective score.

Through block randomization, participants were randomized into two groups (i.e., control and investigational). Both groups received AllerPops as the intervention but with different product consumption instructions. The investigational group was asked to clean their mouth first and let the product slowly dissolve in their mouth for over one hour. The control group was not required to clean their mouth and was instructed to quickly swallow one lozenge every other day for a minimum of 3 doses or until they were satisfied with the symptom relief, up to and including Day 13.

### OUTCOME MEASURES

At baseline and after 7, 14, and 21 days, the TNSS questionnaire was completed by all participants, and PNIF measurements were obtained. Similarly, blood samples were also taken at baseline, day 7, and day 21, to measure total IgE and eosinophils (known immune mediators), and to monitor standard safety parameters. Any out-of-range parameter was flagged and assessed by the Principal Investigator. Heart rate, blood pressure, and body temperature were collected at baseline, day 7, and day 21 to assess safety outcomes. Finally, the emergence, severity, and causality of adverse events were measured on day 7, day 14, and day 21. Saliva samples were obtained from participants at baseline, day 7, and day 21 to evaluate the effect of the AllerPops administration on the oral microbial presentation/microbiota using Amplicon Metagenomics Sequencing. The analysis will be performed separately by Novogene.

### STATISTICS

Continuous data were tested for normal distribution using the Shapiro–Wilk test and were confirmed to be nonparametric. Demographic data was evaluated using mean and standard deviation. Baseline and follow-up values were compared between the investigational and control groups using the Mann-Whitney U test or, where appropriate, Fisher’s exact test. Baseline and follow-up values were also compared within each group, first, over several periods, using repeated measures (RM) ANOVA, followed by the Wilcoxon signed-rank test for each specific period. All analysis was done using RStudio (v. 2022.07.1+554), Microsoft Excel (v. 16.67), and the Real Statistics Resource Pack (Rel 8.3.1).

### 16S PROFILING OF SALIVA SAMPLE

Saliva was collected after fasting for at least 8 hours without stimulation at baseline and after 7, 14, and 21 days and stored at -70 °C immediately. The samples were sent to Novogene (Sacramento, CA) for DNA extraction, 16S amplification, and sequencing. The amplicon sequences were analyzed through Novogene’s pipeline, including quality control, diversity analysis, and abundance comparison. A detailed description can be found in our pilot study[25]. We also did a correlation test between improvements in clinical symptoms score and variation of the OTU abundance, using a function in Microsoft Excel.

## AUTHOR CONTRIBUTIONS

Cliff Shunsheng Han developed the AllerPops allergy theory and wrote the manuscript.

## DATA AVAILABILITY

Sequence data related to this clinical trial is available at Sequence Read Archive, https://www.ncbi.nlm.nih.gov/bioproject/978635, with BioProject ID PRJNA978635.

